# Suicidal Ideation in Dementia: Associations with Neuropsychiatric Symptoms and Subtype Diagnosis

**DOI:** 10.1101/2021.07.10.21253994

**Authors:** Hamish Naismith, Robert Howard, Robert Stewart, Alexandra Pitman, Christoph Mueller

## Abstract

**Objectives:** To investigate factors associated with suicidal ideation around the time of dementia diagnosis. We hypothesised that relatively preserved cognition, co-occurring physical and psychiatric disorders, functional impairments and dementia diagnosis subtype would be associated with higher risk of suicidal ideation.

**Design:** Cross-sectional study using routinely collected electronic mental healthcare records.

**Setting:** National Health Service secondary mental healthcare services in South London, United Kingdom, which serve a population of over 1.36 million residents.

**Participants:** Patients who received a diagnosis of dementia (Alzheimer’s, vascular, mixed Alzheimer’s/vascular or dementia with Lewy bodies) between 1/1/2007-30/6/2016. 11,787 people with dementia were identified during the observation period.

**Measurements:** A natural language processing algorithm was used to identify recorded clinician statements about suicidal ideation around the time of dementia diagnosis.

Sociodemographic and clinical characteristics were also measured around the time of diagnosis. We compared people diagnosed with non-Alzheimer’s dementia to those with Alzheimer’s and used statistical models to adjust for putative confounders.

**Results:** 4.8% of patients identified with dementia had recorded suicidal ideation. Recorded suicidal ideation was almost twice as common in dementia with Lewy bodies compared to other dementia diagnoses studied. After adjusting for sociodemographic and clinical factors, dementia with Lewy bodies, physical illnesses or disability and neuropsychiatric symptoms were significantly associated with suicidal ideation. Mini-Mental State Examination score at diagnosis was not associated with suicidal ideation.

**Conclusions:** Although completed suicide is rare, suicidal ideation is present in sufficient proportions of people with dementia to justify treatment of potentially modifiable risk factors within dementia care pathways.

## Introduction

Around 50 million people worldwide have dementia and this number is predicted to triple by 2050 (Frankish *et al*., 2017; Livingston *et al*., 2017). Disability, dependence, loss of personhood and burden for family members associated with dementia are commonly feared (Haw *et al*., 2009). Although people with dementia are generally at low risk of suicide compared to the age-matched general population, they may be at increased risk in the early period after diagnosis (Haw *et al*., 2009). A recent Danish registry-based study found that whilst the incidence risk ratio in people with dementia was lower overall, risk was elevated in the three months following dementia diagnosis (Erlangsen *et al*., 2020). The overall adjusted incidence risk ratio was 0.8 (95% CI, 0.7-0.9); during the first month after diagnosis it was 3.0 (95% CI, 1.9-4.6; P < .001) (Erlangsen *et al*., 2020). Increased risk of suicide attempt has also been reported in patients with a recent diagnosis of dementia and mild cognitive impairment (Günak *et al*., 2021).

The reasons for these associations and temporal trends are not clear, but several explanations have been proposed. Patients with early dementia might have psychiatric comorbidities, such as depression, which increase suicide risk (Haw *et al*., 2009; Erlangsen *et al*., 2020). Another plausible mechanism for the elevated suicide risk immediately after diagnosis is the distress of receiving a diagnosis (Erlangsen *et al*., 2020; Álvarez Muñoz *et al*., 2020). Initial awareness of the implications of a diagnosis whilst cognition is relatively preserved, and while patients are in possession of the cognitive means to plan and effect suicide, may contribute to this period of increased risk (De Berardis *et al*., 2018; Erlangsen *et al*., 2020).

In light of these findings, we were interested in suicidal ideation (SI) in people with dementia around the time of diagnosis. Previous literature has identified that SI is observed in mild dementia and is strongly associated with depression (Haw *et al*., 2009). However, there are limitations of the current literature, including small sample sizes, unclear definitions of SI and insufficient investigation of the impact of diagnosis subtypes, particularly non-Alzheimer’s dementias (Haw *et al*., 2009; Draper, 2015). We therefore specifically investigated diagnostic subtypes, including vascular dementia and dementia with Lewy Bodies (DLB). It is thought that patients with these disorders may be at increased risk due to the association of these subtypes with neuropsychiatric symptoms such as depression and, especially in the case of DLB, hallucinations and delusions (Haw *et al*., 2009; Armstrong *et al*., 2020).

As the prevalence of dementia rises with increasing age, we were also interested in risk factors for suicidality in older adults, including physical illness, impairment in functioning and activities of daily living, and social isolation (Conwell *et al*., 2011).

We hypothesised that preserved cognitive functioning, co-occurring physical and psychiatric disorders, functional difficulties and some dementia diagnosis subtypes (particularly DLB) would be associated with higher risk of SI.

## Methods

### Data source

Data for this study were collected using the South London and Maudsley NHS Foundation Trust (SLaM) Clinical Record Interactive Search (CRIS) platform. SLaM is one of Europe’s largest healthcare providers for dementia and mental illness and serves a population of over 1.36 million residents across four south London boroughs (Lambeth, Lewisham, Southwark, and Croydon). All services in SLaM have adopted fully-electronic health records since 2006 and CRIS provides research access to over 400,000 anonymized health records within a robust governance framework (Fernandes *et al*., 2013; Perera *et al*., 2016). CRIS has received ethical approval as an anonymized data resource (Oxford Research Ethics Committee C, reference 18/SC/0372) and its functionality is enhanced through application of natural language processing to identified relevant information from free-text records (Perera *et al*., 2016; Mueller *et al*., 2020).

### Study population

We included patients who received a diagnosis of dementia in SLaM services between 1st January 2007 and 30th June 2016. The date of first dementia diagnosis served as index date for analysis. We included patients who received a diagnosis of Alzheimer’s dementia (F00), vascular dementia (F01), a mixed-type dementia (including F00.2 or mentions of both F00 and F01 in the same record) or DLB according to the International Classification of Diseases, version 10 (World Health Organization, 2010). Diagnoses of dementia were ascertained from structured fields, which was supplemented by data from free-text records as previously described (Mueller *et al*., 2020). As there is no unique code for DLB used in mental health records, this diagnosis was purely identified through natural language processing, and the performance of this approach has previously been evaluated (Mueller *et al*., 2018).

### Suicidality

A natural language processing application was used to identify clinician records referring to expressions of SI in a one-year window around the index date of first dementia diagnosis (6 months before or after index date) (CRIS NLP SERVICE, 2020). The algorithm identified SI as the recorded thinking about, considering or planning suicide from text fields in the record. Examples of positive annotations include: ‘Her main concerns were his low mood and ‘the patient’s suicidal ideation’ and ‘He has recently sent a letter (…) describing suicidal ideation’, while examples of negative annotations are ‘There was no immediate risk in relation to self-harm or current suicidal ideation’ and ‘She denies having self-harming or suicidal ideation’. Interrater reliability (Cohen’s k) from wider performance data (not limited to people with dementia), based on 50 documents, was 92%; precision, based on a random sample of 30 patients, was 97% (CRIS NLP SERVICE, 2020).

### Covariates

Sociodemographic factors as recorded at index date were ascertained as follows: age, gender, marital status, ethnicity (dichotomised to White and non-White), and a neighbourhood-level index of deprivation (Noble *et al*., 2007). Cognitive performance was measured through the Mini-Mental State Examination (MMSE) score closest to the initial date of diagnosis (Folstein *et al*., 1975). Co-occurring mental and physical health problems, as well as functional difficulties, were established using the Health of the Nation Outcome Scales (HoNOS65+) (Burns *et al*., 1999). HoNOS65+ is a well-established and validated measure of patient welfare, routinely used in UK dementia and mental services and encompassing twelve clinician-rated subscales. We included scales on difficulties due to neuropsychiatric symptoms, including agitated behaviour, hallucinations and/or delusions or depressed mood, physical illness or disability, and activities of daily living. Each subscale is rated on a scale ranging from 0 (no problem) to 4 (severe or very severe problem). We dichotomised the scores to ‘minor or no problems’ (scores 0 or 1) and ‘mild to severe problems’ (scores 2 to 4) for easy interpretation (Burns *et al*., 1999).

### Statistical Analyses

We used Stata version 15 (StataCorp. 2017, College Station, TX). Descriptive statistics were generated and presented for the full cohort, and whether or not the patient was recorded as displaying suicidal ideation. We applied several logistic regression models to identify cross-sectional predictors of suicidal ideation: first the crude/univariable analysis (model 1) and adjusted for age and gender (model 2), then for all socio-demographic variables and mean MMSE at diagnosis (model 3). Next, further adjustment was undertaken for physical illness (model 4) and additionally for problems with activities of daily living (model 5).

As 22% of the patients included in the final sample had missing data on at least one of the other covariates and we judged missingness in this sample to be random, we imputed missing values using chained equations to maximise statistical power (Oudshoorn *et al*., 1999). Applying the *mi* package in STATA we created 22 imputed datasets through replacing missing values through simulated values assembled from potential covariates and outcome values. Rubin’s rules were applied to combine coefficients in final analyses (Rubin, 2004).

## Results

We identified 11,787 patients diagnosed with the specified dementia subtypes over the observation period. 4.8% had recorded SI. In relation to dementia subtype, 4.4% of patients with Alzheimer’s dementia had suicidal ideation recorded, 4.5% of patients with mixed-type dementia, 5.4% of patients with vascular dementia and 8.8% of those with DLB.

Table 1 compares those with and without suicidal ideation. Patients with suicidal ideation were younger, more likely to be male or from deprived areas and less likely to be married or cohabiting. No difference in MMSE scores was detected between those with and without suicidal ideation. The proportions of patients with DLB and vascular dementia were relatively greater in the patients with suicidal ideation compared to those without. Those with recorded suicidal ideation also had higher levels of neuropsychiatric symptoms and physical illness.

**Table 1:** Sample characteristics for the whole cohort and by the presence of suicidality

| Risk factors | Full cohort<br>(n=11,787) | Patients with suicidal<br>ideation (n=564) | Patients without suicidal<br>ideation (n=11,223) | P<br>value<sup>†</sup> |
| --- | --- | --- | --- | --- |
| **Socio-demographic status and cognitive**<br>**function<sup>‡</sup>** |  |  |  |  |
| Mean age at dementia diagnosis (SD) | 80.6 (9.4) | 75.8 (14.1) | 80.8 (9.0) | <0.001 |
| Female gender (%) | 62.3% | 58.3% | 62.5% | 0.048 |
| Non-White ethnicity (%) | 24.9% | 22.6% | 25.1% | 0.187 |
| Married or cohabiting status (%) | 34.1% | 25.4% | 34.6% | <0.001 |
| Mean index of deprivation (SD) | 27.1 (11.0) | 28.6 (9.9) | 27.1 (11.0) | <0.001 |
| Mean MMSE score at diagnosis (SD) | 18.6 (6.4) | 18.6 (6.5) | 18.6 (6.4) | 0.857 |
| **Dementia subtype** |  |  |  | 0.001 |
| Alzheimer's disease | 46.3% | 42.4% | 46.5% |  |
| Mixed-type dementia (including<br>Alzheimer's disease and Vascular<br>dementia) | 26.3% | 24.7% | 26.4% |  |
| Vascular dementia | 24.6% | 27.8% | 24.4% |  |
| Dementia with Lewy bodies | 2.8% | 5.1% | 2.7% |  |
| **Neuropsychiatric symptoms (%)<sup>§</sup>** |  |  |  |  |
| Agitated behaviour | 19.6% | 30.5% | 19.0% | <0.001 |
| Hallucinations and/or delusions | 13.4% | 23.7% | 12.8% | <0.001 |
| Depressed mood | 15.5% | 37.6% | 14.4% | <0.001 |
| Physical illness or disability (%)<sup>§</sup> | 53.1% | 61.1% | 52.7% | <0.001 |
| Activities of daily living (%)<sup>§</sup> | 59.1% | 62.0% | 58.9% | 0.153 |

† -t-test or Chi2 test; ‡ – at/around the time of dementia diagnosis; § – percentage with HoNOS65+ scores of 2 to 4 at the time closest to dementia diagnosis. Abbreviations: SD, standard deviation.

Table 2 presents logistic regression models for the association of patient characteristics and subtype diagnoses with suicidal ideation. The crude model identified younger age at diagnosis, male gender, not being married or cohabiting and higher area-level deprivation to be associated with greater probability of recorded SI. It also found clinical characteristics of physical illness, all three neuropsychiatric symptoms and vascular dementia and DLB to be associated with greater SI risk. However, in more complex models, the association with the vascular dementia subtype was no longer significant after adjusting for sociodemographic factors, MMSE and physical illness or problems with activities of daily living.

**Table 2:**
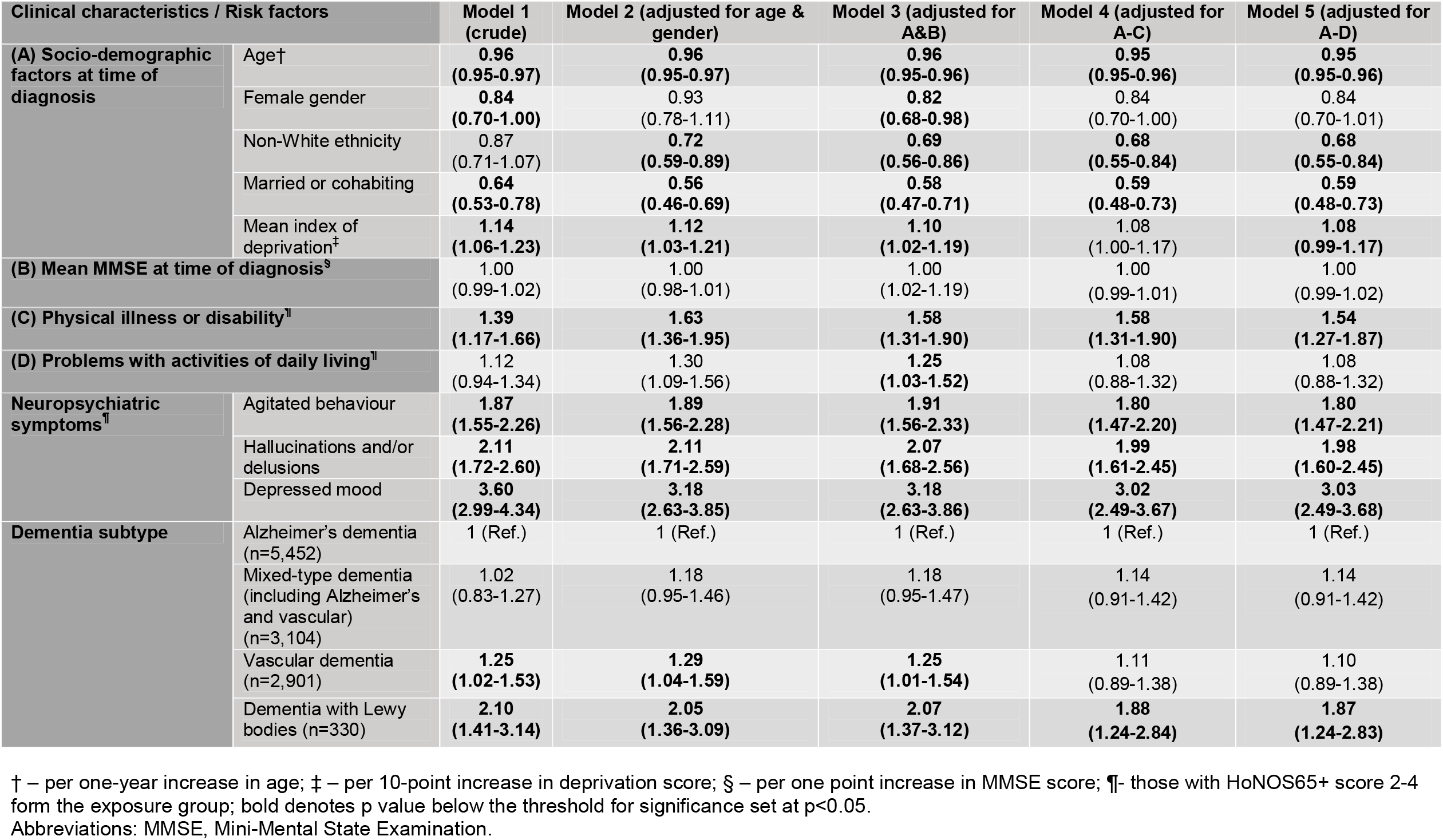
Associations between the characteristics of people with dementia with and without SI and the probability of SI in logistic regression models (presented as odds ratios and 95% confidence intervals)

Our final logistic regression model (adjusted for demographics, MMSE, physical illness and problems with activities of daily living) identified younger age at diagnosis, white ethnicity, not being married or cohabiting and higher area-level deprivation to be associated with greater probability of recorded SI. Clinical factors associated with increased probability of SI included physical co-morbidities, neuropsychiatric symptoms, and DLB sub-type (Odds ratio (OR): 1.87 (95% CI 1.24-2.83)). Depressed mood was the neuropsychiatric symptom associated with the highest risk (OR: 3.03 (95% CI 2.49-3.68)) in the final model. Gender, mean MMSE at diagnosis, activities of daily living and diagnosis of mixed-typed dementia or vascular dementia were not associated with increased risk in the final model.

## Discussion

In our study of more than 11,000 patients in routine health care, 4.8% of people with dementia had a record of suicidal ideation around the time of diagnosis. After adjusting for sociodemographic and clinical factors, significant associations were detected between SI and DLB, physical illnesses or disability and neuropsychiatric symptoms, while cognition measured through MMSE score at diagnosis was not associated with suicidal ideation.

Draper (2015) noted that few previous studies have reported prevalence of suicidal ideation but our data can be compared to an Australian study of outpatients with dementia by the same author. The study reported the prevalence of different severities of SI using the Hamilton Depression Rating Scale criteria: 5.4% of patients felt life was not worth living, 3.2% reported a “ wish to die” and 0.9% reported suicidal ideation or gestures (Draper *et al*., 1998).

We found younger age was associated with SI, which is consonant with other studies of suicidality in people with dementia and may reflect difficulties in adjusting to the diagnosis (Draper, 2015; Álvarez Muñoz *et al*., 2020). Other predictors identified are consistent with known risk factors in older adults without dementia. We found physical illness or disability to be associated with greater risk of SI; in older adults, greater numbers of comorbidities contributes to cumulative suicide risk (Conwell *et al*., 2011). We found patients who were not married or co-habiting had a greater risk of SI; loneliness, low social support and loss of a spouse have also been associated with increased risk in older adults (Conwell *et al*., 2011).

As noted above, the preservation of executive function may be plausibly associated with the development of SI. The lack of association between SI and the MMSE in this study may reflect the relative insensitivity of the MMSE to impairments in executive functioning (Malloy *et al*., 1997).

Neuropsychiatric symptoms were significantly associated with SI, in particular depression. This is in keeping with previous research showing depression is a risk factor for SI in dementia and for suicide in later life (Conwell *et al*., 2011).

After confounder adjustment patients with DLB were at 1.87 higher odds of suicidal ideation than those with Alzheimer’s dementia. There are several possible explanations for this. First, patients with DLB might have a greater burden of non-cognitive symptoms, which significantly impact quality of life (van de Beek *et al*., 2019). These include delusions and hallucinations, behavioural disturbances and sleep disorders (Galvin *et al*., 2010; Armstrong *et al*., 2020). Second, patients with DLB may have fluctuating insight into their cognitive limitations and times of greater insight and functioning could plausibly be associated with increased risk (Armstrong *et al*., 2020). Finally, patients and their carers report negative experiences of the diagnostic process with DLB: they may attend multiple consultations over years and receive a delayed diagnosis or an initial misdiagnosis (Galvin *et al*., 2010; Armstrong *et al*., 2020; Surendranathan *et al*., 2020).

Vascular dementia subtype was associated with increased risk in the unadjusted model, but notably this association was no longer significant in more complex adjusted models, such as those incorporating physical illness. This may be because patients with vascular dementia are more likely to have comorbidities such as diagnoses of stroke (Pendlebury *et al*., 2009), which themselves are associated with increased risk of suicide (Erlangsen *et al*., 2020).

### Strengths and limitations

Strengths of this study are the large sample size compared to previous studies: to our knowledge, this is the largest cross-sectional study of dementia and suicidal ideation to date (Álvarez Muñoz *et al*., 2020). A further strength is the specific comparison of non-Alzheimer’s dementias to Alzheimer’s. Our use of routinely collected data minimises response bias and we incorporated widely used and validated measures (MMSE and HONOS65+).

Limitations include a reliance on the accuracy of clinical data entries and on the natural language processing algorithm to bolster the detection of covariates. The algorithm for SI detection was not limited to people with dementia. As such, internal validity may be affected if people with dementia express SI differently to other patients in secondary mental healthcare settings and further investigation of this should be undertaken. We also acknowledge that we would not have detected SI if clinicians had not enquired, or patients declined to disclose it.

### Conclusions and future research directions

The association of neuropsychiatric symptoms, physical illness or disability and DLB subtype with greater probability of SI highlights the need for further research to delineate potential causal relationships between the studied covariates and suicidality. This could be done using longitudinal designs and including more detailed measures of executive function in the months and years after diagnosis. Additional research in other locations would also inform the external validity of this study’s findings.

Further study of the associations between SI, suicide attempt and suicide would be of interest to clinicians in understanding the risk of these different outcomes. The potential effect of addressing modifiable risk factors such as depression and other neuropsychiatric symptoms should be undertaken. The potential for post-diagnostic support to reduce suicidality should also be investigated; the introduction of community support for people with dementia is one suggested explanation for a lower suicide rate in more recent cohorts of people with dementia in Denmark (Erlangsen *et al*., 2020).

Efforts to reduce the estimated 4.8% of patients expressing SI after a dementia diagnosis may be served by further training memory clinic staff to ask about SI and addressing modifiable risk factors. Involving carers in medication safeguarding as part of means restriction may reduce risk as the suicide prevention intervention supported by the strongest evidence (Zalsman *et al*., 2016).

## Data Availability

The data accessed by CRIS remain within a National Health Service firewall and governance is provided by a patient-led oversight committee. Subject to these conditions, data access is encouraged and those interested should contact R.Stewart, CRIS academic lead.

## Description of authors’ roles

C. Mueller, R. Howard and H. Naismith formulated the research question; C. Mueller ran the CRIS data extraction and statistical analysis; C. Mueller and H. Naismith wrote the first draft of the manuscript; all authors read, commented on, and contributed to the final manuscript.

C. Mueller and R. Stewart are part-funded by the National Institute for Health Research (NIHR) Biomedical Research Centre at South London and Maudsley NHS Foundation Trust and King’s College London, which also supports the development and maintenance of the Biomedical Research Centre Case Register. The views expressed are those of the author(s) and not necessarily those of the National Health Service, the NIHR or the Department of Health and Social Care.

## Acknowledgements

The authors wish to thank the Clinical Record Interactive Search (CRIS) team. The data accessed by CRIS remain within a National Health Service firewall and governance is provided by a patient-led oversight committee. Subject to these conditions, data access is encouraged and those interested should contact R. Stewart, CRIS academic lead.

